# Perspectives of Researchers and Clinicians on Patient and Public Involvement (PPI) in Preclinical Spinal Cord Research: An Interview Study

**DOI:** 10.1101/2023.08.29.23294719

**Authors:** Pádraig Carroll, Adrian Dervan, Ciarán McCarthy, Cliff Beirne, Geoff Harte, Dónal O’Flynn, John Quinlan, Éimear Smith, Frank Moriarty, Fergal J. O’Brien, Michelle Flood

**Affiliations:** School of Pharmacy and Biomolecular Science, Royal College of Surgeons in Ireland University of Medicine and Health Sciences, Dublin, Ireland; Tissue Engineering Research Group (TERG), Department of Anatomy and Regenerative Medicine, Royal College of Surgeons in Ireland University of Medicine and Health Sciences, Dublin, Ireland; Advanced Materials and BioEngineering Research (AMBER) Centre, Trinity College Dublin (TCD) and RCSI, Dublin, Ireland; c/o Irish Rugy Football Union Charitable Trust, Dublin, Ireland; Faculty of Sports and Exercise Medicine (Royal College of Physicians in Ireland & RCSI), Dublin, Ireland; Tallaght University Hospital, Tallaght, Dublin, Ireland; National Rehabilitation Hospital, Dublin, Ireland; PPI Ignite Network, Galway, Ireland

## Abstract

**Study Design:** Qualitative study

**Objective:** To explore the perspectives of preclinical spinal cord researchers and clinicians involved in the treatment of spinal cord injury on patient and public involvement (PPI) in preclinical research.

**Setting:** Preclinical spinal cord injury research.

**Methods:** Semi-structured interviews were conducted online to collect data that was analysed thematically.

**Findings:** Twenty-two participants (11 clinicians and 11 preclinical researchers) were interviewed. Participants recognised the value of PPI in improving the relevance of preclinical spinal cord research and providing a source of motivation for lab-based research. The perceived distance between preclinical research and the day-to-day experiences of PPI contributors was identified as a major barrier. Inclusive practices and the highly networked and motivated community of people affected by spinal cord injury were noted as facilitators. Building strong partnerships was considered essential for successful PPI.

**Conclusions:** While PPI has traditionally been more commonly associated with clinical research, participants identified the potential benefits of PPI in preclinical spinal cord research to provide context and improve research relevance and impact. Preclinical researchers should explore how PPI can be incorporated in their work.

## Introduction

Patient and Public Involvement (PPI) is the active and meaningful engagement of patients and/or the public in the research process ^1^. It is more commonly referred to as patient engagement in North America. At its core, PPI appreciates the experience and perspectives of those living with particular conditions in shaping and improving research and considers those who will be most affected by the results to have the right to an opportunity to input ^2, 3^. PPI can involve patients and/or the public at any stage of the research process, for example identifying research priorities, or informing dissemination strategies to share results with wider audiences ^1^. It can take many forms from patients and the public sitting on steering groups or committees to collaborating as co-researchers ^4^. A growing evidence base underpins the benefits of PPI in enhancing research quality and appropriateness, enhancing clinical study recruitment strategies, and improving the dissemination of findings ^2^.

In spinal cord injury (SCI) research, there is increasing recognition that people living with spinal cord injury should be meaningfully involved as partners across all stages of the research process with appropriate remuneration ^5, 6, 7^. Examples of successful partnerships in spinal cord research include the development of a set of research priorities for spinal cord injury ^8^. However, the field of spinal cord research spans preclinical (including basic, fundamental, biomedical, translational, or lab-based research) and clinical (including rehabilitation, occupational, clinical trials) settings and it is important to consider how involving patients and the public can be optimised for these diverse contexts. While the evidence base for PPI in clinical research is well established and has become a recognised and important feature of research practice for key stakeholders including funding agencies, regulators, and journal editors, the impact off PPI in the field of preclinical research has not been as comprehensively explored.

For PPI in preclinical research, the technical nature, need for specialist knowledge, and limited access to the laboratory environment has made it a less obvious setting for PPI ^9^. This is reflected in the limited empirical evidence base for PPI in preclinical research identified in two recent reviews ^10, 11^. The available literature indicates that researchers’ perspectives on preclinical PPI reported vary, with some authors suggesting PPI can reduce waste, increase value, and improve the quality of preclinical research, but others caution that it is difficult to meaningfully involve patients and the public in this context ^9, 12^. Combined, the limited evidence base to underpin preclinical PPI and diverse perspectives reported there make it challenging for preclinical spinal cord research teams to find guidance on getting started. Due to the limited guidance on PPI for preclinical research ^10,11,12^, and the importance of research participation to the SCI community ^13^, it is timely to explore the role of PPI in preclinical SCI research.

Our PPI Advisory Panel highlighted this challenge when developing a strategy for our PPI work for a preclinical spinal cord repair regenerative medicine project. The study is a collaboration between the Irish Rugby Football Union Charitable Trust (IRFU CT), a charity that supports rugby players living with SCI and the Science Foundation Ireland Advanced Materials for BioEngineering Research (SFI AMBER) Centre located at the RCSI University of Medicine and Health Sciences and Trinity College Dublin. This ambitious multi-disciplinary project aims to develop an advanced scaffold-based platform combining stem cells, gene therapy and electrostimulation aimed at providing a solution for spinal cord repair ^14,15^, and is overseen by a PPI Advisory Panel consisting of preclinical researchers, clinicians, and people living with rugby-related spinal cord injury to help guide the research. As the PPI Advisory Panel became more established it quickly became apparent that there was limited guidance available form the published literature, and a secondary aim to contribute towards an evidence base for PPI in preclinical spinal cord research was agreed. Since researchers and clinicians work closely with patients and the public as part of PPI activities, we were interested in their perspectives and how this might influence preclinical spinal cord PPI.

The aim of this study was thus to explore the perspectives of preclinical spinal cord researchers and clinicians involved in the treatment of spinal cord injury on PPI in preclinical research. It aimed to understand their perspectives on what the role and goals of PPI should be in preclinical spinal cord research and their views on potential benefits and barriers to gain a better understanding of PPI in this context.

## Methods

### Design

This qualitative study used semi-structured interviews to gather data from participants. This method combined a pre-determined set of questions (see Table 1) with the flexibility to explore particular responses further if needed. The Human Research Ethics Committee (REC) at RCSI University of Medicine and Health Sciences provided ethical approval for the study (RCSI REC Record ID: 212494891). All participants provided written informed consent before participating in the study.

**Table 1.**
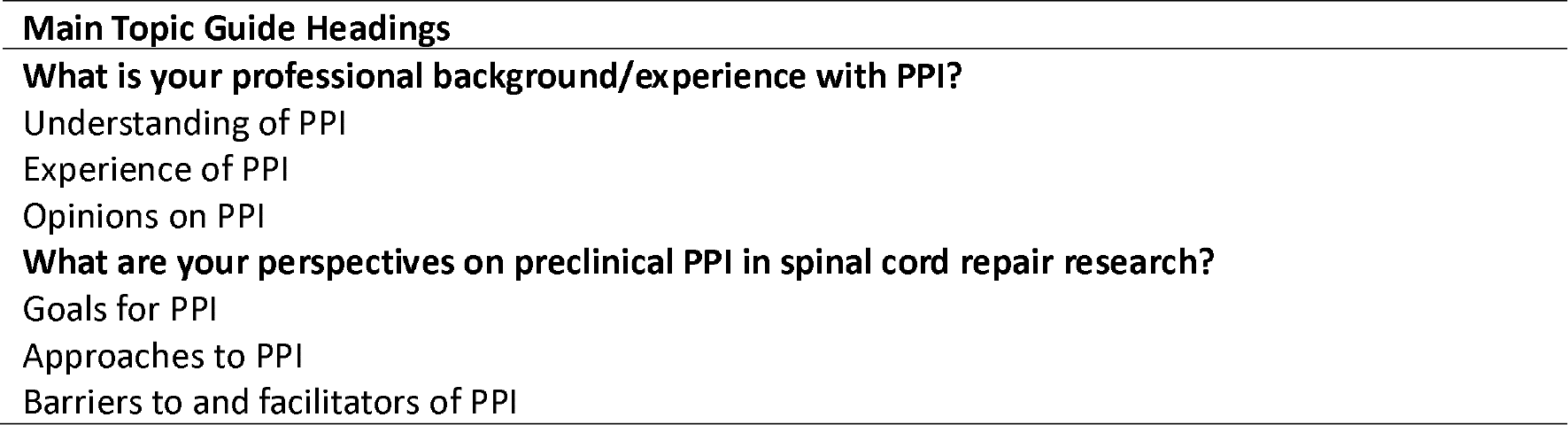
Topic Guide Summary.

### Participants

Preclinical spinal cord researchers and clinicians involved in the treatment of spinal cord injury were eligible to participate. In this study, ‘preclinical researchers’ refers to research scientists working primarily in laboratory settings, and ‘clinicians’ refers to qualified medical doctors. Researchers and clinicians at all stages of training/practice were eligible to take part. Participants were excluded if they had less than six months of experience working in their respective fields.

### Sampling and recruitment

Purposive sampling was used to identify eligible participants. For preclinical researchers, potential participants were first identified by their publishing record in the field of SCI and contacted via email. The introductory email contained an information leaflet, consent form, and contact details for the lead author of the current study (PC). PIs were asked to circulate the information leaflet to members of their research groups, who then contacted PC to arrange an interview if interested. Clinicians were contacted and recruited using a similar process. The research study was also advertised using social media sites including Twitter and LinkedIn and the professional and personal networks of the research team. Participants were offered a €30 gift voucher in recognition of their time for participation. The study sample size was guided using the concept of information power to determine the number of participants needed ^16^. Due to the narrow study aim, specificity of the study to spinal cord injury, and experience of the interviewer, the aim to recruit approximately 10 participants for each group was agreed as a goal by team members ^16^.

### Data Collection

The study PPI Advisory Panel provided feedback on questions and terminology and approved the interview guide prior to commencement of the interviews and collection of data. An abridged topic guide is included in Table 1. Participants were asked initial demographic questions, including information about their current role and any previous experience with PPI followed by open ended questions relating to their perspectives on PPI in preclinical spinal cord research. Interviews were conducted by PC, a male PhD student with formal training and experience in qualitative research and interviewing and took place from March to May 2023. Interviews were conducted and recorded using Microsoft Teams and then transcribed verbatim by PC.

### Data Analysis

All recorded and transcribed data was analysed inductively using the framework described by Braun and Clarke for thematic analysis (TA) ^17^. PC met regularly with MF during the analysis process to discuss progress and challenges. Themes were generated using the following six phases – familiarisation, coding, generating initial themes, reviewing and developing themes, refining themes, and writing up ^17^. Final themes were grouped under three broad headings (1) perspectives on preclinical PPI, (2) barriers and enablers, and (3) developing partnerships in PPI. The findings from this analysis are summarised in Figure 1. The COnsolidated criteria for REporting Qualitative research (COREQ) checklist was used to guide study reporting ^18^.

**Figure 1.**
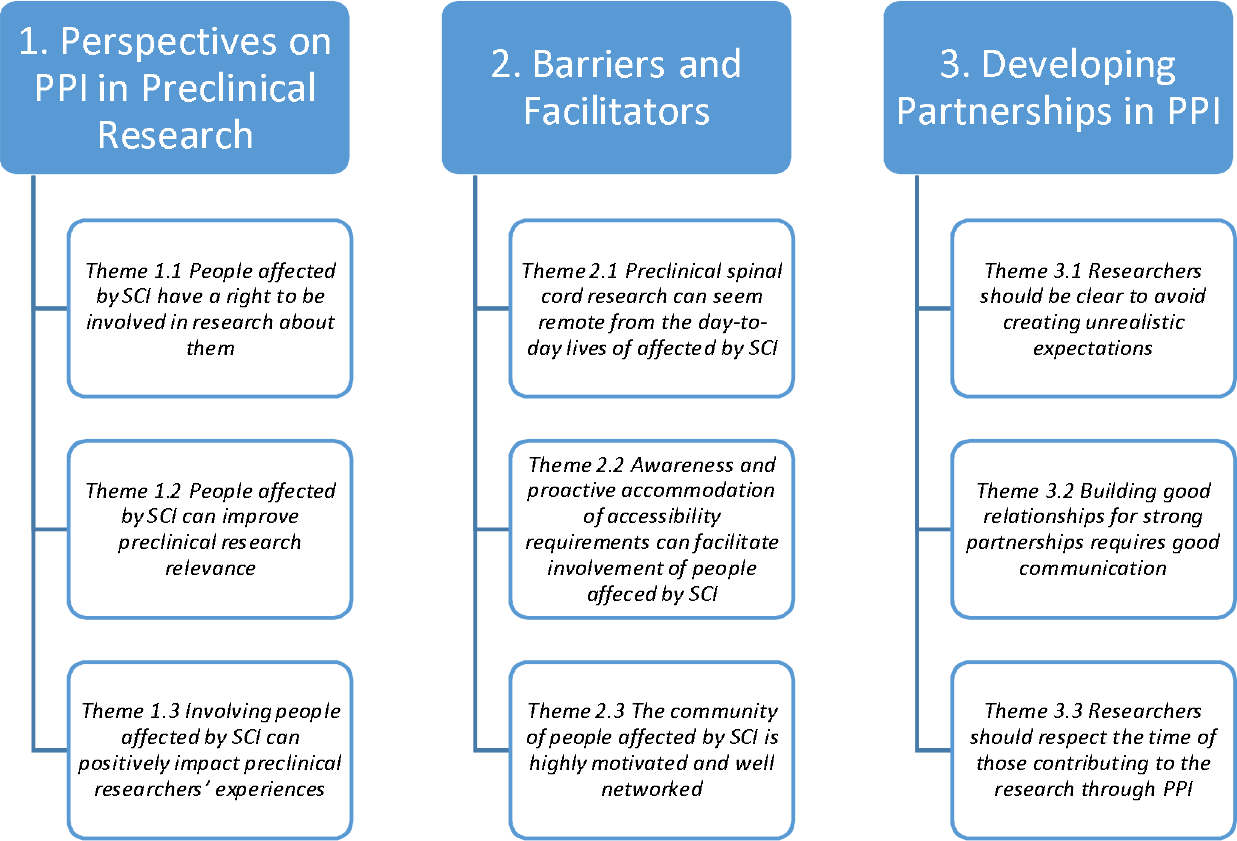
Summary of Themes from Analysis

## Findings

### Demographics

There were twenty-two participants in total, eleven preclinical researchers and eleven clinicians. All those who contacted PC and were eligible to participate were interviewed so eleven were interviewed in each group, which aligned with the recruitment aim. Among the researchers, there were two principal investigators, one research fellow, two postdoctoral researchers, and six PhD students. Among the clinicians, there were seven consultants, one specialist registrar, and three registrars. The specialities represented in this group included orthopaedic surgery, neurology, intensive care medicine, and rehabilitation medicine. Nineteen participants were based in the Republic of Ireland; the other three were based in the USA, Singapore, and the United Kingdom. All participants were familiar with the concept of PPI, with nine having previous experience of participating in a PPI activity. Participant demographics are summarised in Table 2.

**Table 2.** Participant Demographics.

| **Interview ID** | **Group** | **Gender** | **Professional Role** |
| --- | --- | --- | --- |
| **R1** | Researcher | F | Principal Investigator |
| **R2** | Researcher | F | PhD Student |
| **R3** | Researcher | F | PhD Student |
| **R4** | Researcher | F | Principal Investigator |
| **R5** | Researcher | F | Postdoctoral Researcher |
| **R6** | Researcher | M | PhD Student |
| **R7** | Researcher | M | PhD Student |
| **R8** | Researcher | M | PhD Student |
| **R9** | Researcher | M | Postdoctoral Researcher |
| **R10** | Researcher | F | PhD Student |
| **R11** | Researcher | M | Research Fellow |
| **C1** | Clinician | F | Specialist Registrar in Rehabilitation Medicine |
| **C2** | Clinician | F | Consultant in Intensive Care Medicine |
| **C3** | Clinician | F | Consultant Orthopaedic and Spine Surgeon |
| **C4** | Clinician | M | Consultant Orthopaedic and Spine Surgeon |
| **C5** | Clinician | M | Registrar in Rehabilitation Medicine |
| **C6** | Clinician | F | Registrar in Neurology |
| **C7** | Clinician | M | Consultant Orthopaedic and Spine Surgeon |
| **C8** | Clinician | M | Registrar in Rehabilitation Medicine |
| **C9** | Clinician | F | Consultant in Rehabilitation Medicine |
| **C10** | Clinician | F | Consultant in Rehabilitation Medicine |
| **C11** | Clinician | M | Consultant Orthopaedic and Spine Surgeon |

### Themes

The themes generated from the analysis are presented in Figure 1 grouped under three subheadings for clarity.

### 1. Perspectives on PPI in Preclinical Research

Clinician and researcher participants reported having awareness of PPI as a concept, but their perspectives varied based on their level of experience with PPI and professional backgrounds. The clinicians tended to relate their understanding of PPI to their clinical experience in healthcare e.g. *“So, kind of involving patients as partners in their own healthcare. And I guess it’s kind of a similar concept being carried over to research that we should involve the people”* (Clinician 1) and researchers described learning about PPI as part of their research practice through attending conferences or writing grant funding applications. For a smaller number, they had direct experience with clinical PPI activities.

*Theme 1*.*1 People affected by SCI have a right to be involved in research about them*

Both researchers and clinicians expressed support for PPI in preclinical spinal cord research based on the belief that people have a right to be involved in research that could directly impact their lives. PPI was considered important since public money frequently funds research: *“The public should be aware of what we’re doing because their money has been used to pay for us”* (Researcher 8) and since available funding is limited, it is important to ensure research is focused on achieving important patient goals rather than purely academic interests:

> *“In terms of things like spinal cord injuries or where you’ve got a much more distinct patient voice, it’s more specific, and I guess if there is money being spent on research for a certain condition, we should definitely be asking them what they think that money would be best spent on, or what they would like the focus of that should be, what outcomes they would like to see achieved, not just doing things for the sake of doing them and for academia’s sake*” (Clinician 5).

*Theme 1*.*2 People affected by SCI can improve preclinical research relevance*

The majority of participants identified that PPI could improve the relevance and focus of preclinical research and prevent time being spent on ideas that would not have practical application:

> *“If you only had this PPI thing at the clinical stages of work, then your research effectively builds a product that you may or may not be able to apply as opposed to if you start with clinicians and patients early, you know exactly what the problem is because you’ve asked the patients about the problem, you’re not assuming the problem, and if you talk to clinicians, you think about what’s applicable and what’s not”* (Researcher 11).

PPI was also considered to help reduce the risk of preclinical research projects failing to translate their outputs into clinical application: *“A lot of the time in biomedical research, that next step doesn’t happen, and biomedical research start-ups have like a 95% failure rate”* (Researcher 9). Clinicians highlighted the role of PPI in identifying functional goals with importance to those living with SCI: *“What’s important to the patients is not always as important to the clinician. So an increase in the half dozen points of your ASIA score is nowhere near as important as improving your bladder control or giving you functional hand activity so you can feed yourself*.*”* (Clinician 4) and also noted the importance of involving clinicians as well as patients and the public:

> *“Obviously, spinal cord injury is not exactly binary. This huge amount of physical and psychological and social aspects of spinal cord injury, and there’s no kind of size fits all score or goal that you want to try to achieve. So, who (do) you want even involved in assessing what’s important? And then also it’s a very high-risk group as well. So it needs a lot of research. So identifying what the most important things for patients are for those reasons is obviously really important to make sure that you’re actually assessing and tackling issues that patients are concerned about or that the patients and doctors are concerned about and then obviously then you’re measuring them in a way that’s actually as close to the real world as possible. And I think involving patients and doctors and their carers. Not just doctors, but like all the healthcare professionals looking after patients in spinal cord injury, it would be important that way*.*”* (Clinician 8)

Some participants acknowledged that PPI contributors may not have the technical expertise required to influence preclinical research: *“Obviously if they’re not from a scientific background themselves, then their understanding of scientific principles may limit the area of research that can be explored”* (Clinician 7). They noted their input may not directly affect day-to-day experimental research: *“It’s*

*(PPI) not going to impact my experiments”* (Researcher 3), *through the sharing of their experiences with researchers, they can still guide them to address particularly important challenges:*

> *“I think the most useful, and best thing that can be done in the fundamental research, I would try to really understand the symptoms of spinal cord injury and what the patients’ experiences are and then try to understand the kind of pathologies that are causing those symptoms in the patients”* (Researcher 9).

*Theme 1.3 Involving people affected by spinal cord injury can positively impact preclinical researchers’ experiences*

PPI was noted to have benefits for preclinical researchers themselves, for example by helping them retain focus on why they are doing the research and keeping in mind the long-term goals of helping those affected by spinal cord injury:

> *“From the standpoint of someone who’s always worked in a lab setting, not always in spinal cord injury, but at least for the last four years in spinal cord injury, it’s very easy for us to get distracted maybe? Because our focus is to get very much into the details and the nitty-gritty of how we’re going to run experiments”* (Researcher 5)
>
> *“There’s a very real element that is very present when you’re doing PPI, that can get lost at other times just by the nature of the job being occasionally quite far away, especially in preclinical research. So to be reminded of the fact that there are people who you will eventually be doing this for is nice. Yeah reminds me why I got into it”* (Researcher 6)
>
> PPI was also reported to serve as a source of motivation for researchers in the field: “I feel like it brings a lot of benefit to the researchers, in terms of motivating them….and for the patients who were involved, for hope and directing the outcomes” (Researcher 6).

### 2. Barriers to and Facilitators of PPI in Preclinical Spinal Cord Research

Participants identified barriers to and enablers, for PPI in preclinical spinal cord research relating to conceptual and practical considerations.

*Theme 2*.*1 Preclinical spinal cord research might seem remote from the day-to-day lives of those affected by SCI*

Several participants referenced the main challenge of PPI in preclinical spinal cord research being that preclinical research does not immediately benefit those living with SCI. They suggested that this may make it more difficult to attract people to be involved: *“I guess it’s (research outputs) not of immediate benefit to patients with existing spinal cord injury. You’re more pitching it as something that will benefit future people. So it’s not of direct benefit to* the *people you’ll be trying to recruit*.*”* (Clinician 1). In addition, the amount of time it takes to successfully translate preclinical research may not be commonly understood:

> *“Research is slow. It’s the nature of research, approvals, ethics, everything, everything always takes time, but we’re talking about the time like it’s years and years, not like days, or months. This is something that takes up a long period of time and when you oversimplify it, you believe it is ingrained in the fact that you might think it will take a few months, and then after that, if that keeps getting repeated and repeated and repeated, then you don’t really see that light at the end of the tunnel”* (Researcher 11)

*Theme 2*.*2 Awareness and proactive accommodation of accessibility requirements can facilitate involvement of people affected by SCI*

Participants noted that people living with SCI might have additional requirements including accessible transport and facilities that should be considered when organising PPI activities. Accessible facilities and providing hybrid or online options were considered facilitators. When planning preclinical PPI activities for those with spinal cord injury, such considerations were considered important for success.

> *“One would be like a physical barrier, depending on their injury and you know whether they can easily use transport. I know you can do it over Zoom but depending on the severity of their condition, virtual meetings would help with that. I mean I don’t know, maybe some people are just not really interested in research”* (Researcher 2)
>
> *“Our lab is designed to be ADA compliant, so there are benches for people who use wheelchairs. And so I think that really helps to have the infrastructure in place to welcome people with spinal cord injury into your lab”* (Researcher 5)

*Theme 2*.*3 The community of people affected by spinal cord injury is highly motivated and well networked*

Several clinicians explained that from their experience that people living with spinal cord injury are motivated and familiar with and interested in research: “*My experience of many people with spinal injuries, [they] are very motivated. They’re very well-read around the whole area, so inviting them to be involved in the whole process would be very beneficial”* (Clinician 11), *“Some of my patients are quite academic, and they would love to have a one-to-one with somebody to bounce ideas off”* (Clinician 9). Others noted that they are well networked, and communicate well and so should be a facilitator for PPI:

> *“Another interesting thing with cord injured patients, is they’re a small group of people and they’re quite well networked. So informing a large group of patients can happen easily through a small number of people who are involved in the research because they do have that kind of well-established network in place*.*”* (Clinician 4)
>
> *“And so using people like [spinal injury charity] and also [Rehabilitation Centre] to maybe promote research and sort of explain how patients can be involved at various different (stages), not only as participants but also, as you know, contributors”* (Clinician 9)

## 3. Developing Partnerships in PPI

Participants noted a number of factors were important for building strong partnerships with people affected by spinal cord injury in PPI. Being clear and proportionate about potential research outcomes and challenges to avoid creating unrealistic expectations, respecting people’s time, and communicating well were considered important by participants.

*Theme 3*.*1 Researchers should be clear and realistic to avoid creating unrealistic expectations*

A common concern among participants was the potential for PPI to create unrealistic expectations and disappointment or disillusionment: *“The more studies to come through promising something to cure XYZ to improve outcomes that don’t end up being anything……I suppose they (people with SCI) would be quite jaded after hearing about a lot of our research that doesn’t come to fruition”* (Researcher 6). They felt that this was particularly relevant for preclinical research given the lengthy timelines involved in completing a research project *“Say if you work in research or you’re working in healthcare, you might understand that a pre-clinical trial if it works, could be decades away from having an impact on actual clinical outcomes and I guess that would be the one bit in my mind, I’d want to be a little careful”* (Clinician 5). Having a clear sense of the aim of PPI and how it would benefit people involved to prevent the development of ‘false hopes’ and being honest were considered important:

> *“I just would be very, very reticent and very keen to protect patients from having unreal expectations because you know there’s going to be a huge body of work between the doctors, the psychologists, the physiotherapists to support them[people with SCI] in a way that is kind and that is realistic and you know, you can understand how someone like that would be so vulnerable if someone came and said to them, “well, I want to involve you in research” and you know, like that it might engender false hopes. So I guess it would be really important that if you’re involving patients that you know, you were very clear about what the hope and the aim are and would it benefit them personally”* (Clinician 2)
>
> *“I suppose that the patient can bring their perspective on how you know what somebody’s proposing to do might change somebody’s life. It depends because some of these things that I imagine you’re (researchers) doing won’t help every patient with spinal cord injury and it might be future patients or a certain cohort of patients, and there might be lots of inclusion and exclusion criteria and people have to understand that. But I think if you’re honest with them and don’t sort of sugar-coat it as though this is the miracle cure…*.*I think always being honest is key you know*.*”* (Clinician 10)

*Theme 3*.*2 Building good relationships for strong PPI partnerships requires time and honest communication*

Participants identified how good communication leads to the development of rapport which is important for PPI:

> *“I think that [rapport] will only come after time because people aren’t generally, with some exceptions, going to have the confidence to say that in the first meeting. And so I think that that’s why relationship building is so important to be like when we do the PPI work now we get great questions about it and very searching questions as both their understanding of our work grows and the relationship gets better as well”* (Researcher 6)

They also emphasised the importance of aspects such as feedback, and being clear, open, and honest, including about challenges faced by preclinical researchers:

> *“So I suppose, you know, everyone would be capable of understanding. You don’t have to have a degree in science or whatever to maybe be able to understand what’s done in the lab. The idea is that it’ll translate into meaningful treatment for either them or other patients or future patients. And so I think just involving them, being open, putting it in terms that they can understand, they don’t have to understand everything to understand”* (Clinician 10)
>
> *“So for patients to realise that we are still trying to find ways to repair cords because we don’t really have one at the moment unfortunately. So they could have to feel that we are still trying to find ways to push the technology forwards from that point of view. I think it’s very important for them to see the difficulties that go with how we move through research as well”* (Clinician 3)

*Theme 3*.*3 Researchers should respect the time of those contributing to the research through PPI*

Participants noted it was important that any processes would not place too much of a demand on participants. Others noted that reimbursing participants for their time would be appropriate if possible.

> *“There needs to be some mechanism for ongoing feedback in both directions where researchers continuously update patients but not in a way that’s too burdensome for them to be effectively engaged and for research to also get feedback from the patients and what the patients would like to see done, especially as we move towards stuff that’s a bit more pre-clinical than fundamental”* (Researcher 9)
>
> *“There probably isn’t enough money to pay people for their time, but it would make it better. And I could imagine people would have very good engagement”* (Clinician 8)

## Discussion

There is limited evidence on the role of PPI in preclinical research when compared with PPI in clinical research. This study aimed to explore the perspectives of preclinical researchers and clinicians on PPI in preclinical spinal cord research. Both groups are key stakeholders in PPI and therefore understanding their perspectives is important. The findings from this study suggest that for the preclinical researchers and clinicians interviewed, there is an understanding of the potential benefits of PPI in preclinical research. They note its usefulness for the research and researchers, can identify barriers and facilitators, and appreciate the importance of building strong relationships with patients and the public who are involved. They recognise the role of people affected by SCI as experts that bring their knowledge based on their lived experience.

The themes relating to participants’ perspectives map broadly to values associated with PPI in health and social care research: substantive values (including quality and relevance), normative values (ethical/political issues relating to empowerment), and process values (including partnership, honesty, and clarity) ^19^. The participants, both researchers and clinicians, suggested that PPI can benefit preclinical spinal cord research and that it could be particularly useful in identifying research priorities and ensuring that limited funding is used optimally where it could have most impact. This viewpoint reflects the wider emphasis on identifying research priorities in the preclinical PPI literature when compared with other activities such as collecting and analysing data, which has been reported in clinical research ^10, 11^. Other studies have suggested that involving individuals living with spinal cord injury at the earliest stages of research can help prevent failure of research to translate ^20^. Participants also identified that for preclinical spinal cord researchers, PPI may serve as an important source of motivation through reminding them of the bigger picture and their ultimate goal. This viewpoint is related to process values associated with health research but takes a different form in preclinical research, where interaction with patients or the public is not as common. Some also regarded those affected by SCI to have a moral or ethical right to input to research that might ultimately, even though not directly, impact them as clinical research would be needed before any of the preclinical research findings could impact patient care (normative values). Overall, the perspectives of participants reflected those associated with health and social care researchers, with slightly different emphasis on aspects such as being a source of motivation for laboratory-based researchers that are not generally associated with PPI in clinical research. This may benefit from further study to fully understand its impact in preclinical spinal cord research and the laboratory research community more broadly.

In terms of barriers and enablers, participants identified the primary barrier as being that preclinical research typically happens in laboratories and outputs that do not have immediate practical application for patient care, and that while successful outputs may happen, they require a long time and significant further research including clinical research. They understood this to be a particular challenge not encountered when PPI forms part of clinical research. Other studies have also referred to this gap as being potentially problematic ^21, 22^. However, there are an increasing number of studies reporting the process and impact of PPI in preclinical research, and despite the concerns of the preclinical researchers and clinicians it appears that with appropriate preparation and support it can be achieved ^10, 11.^ Proactively managing accessibility considerations reflects an important practical consideration to ensure that PPI contributors’ requirements are anticipated and met ^23^. Finally, the highly motivated and networked group of people affected by SCI were identified by several participants serves as an important facilitator and finding ways to connect with patient groups and individual patients can help researchers identify people willing to participate in PPI activities. This is encouraging for preclinical researchers.

To develop strong partnerships for preclinical PPI in spinal cord research, researchers should be mindful of communicating clear and realistic expectations, dedicating time and being honest, and respecting the time of PPI contributors. Clinicians emphasised that it would be important to clearly communicate the preliminary nature of research findings, as they believed people with SCI might develop high expectations for a cure from early research findings. This contrasts with views expressed by people with SCI, who reported that being involved in PPI could have the opposite effect, that learning more about research would reduce their hope for a cure ^24^. The importance of developing strong relationships noted by study participants is generally accepted to be essential for successful PPI but can be challenging to achieve ^23, 25^, and has formed the basis of a model for PPI in SCI ^13^. Previous research on PPI in preclinical research indicated that researchers may feel that they lack the skills to communicate effectively with patients or patient organisations, however this was not a primary concern identified in this study ^21^. It is possible that the focus on communication skills as a core skill for all researchers has reduced this concern. Previous research also suggests that PPI improves researchers abilities to communicate their findings to the wider public ^26, 27^. Communication includes providing feedback to PPI contributors about research outputs from preclinical researchers ^28^, but this was not mentioned by the study participants, which may highlight a need for this to be included in training sessions. Provision of feedback on the results of scientific studies may appear more challenging initially, but our own experience of including updates of research progress and findings has been positive. Another important consideration is providing payment for PPI contributors’ time and expenses, which is increasingly considered to be essential and has led to the development of specific guidelines for researchers ^7^.

This study adds to the small but growing empirical evidence base for PPI in preclinical spinal cord research ^10,11^. Our findings are relevant considering other studies, suggesting that PPI provides an alternative form of knowledge to preclinical research ^29^, connects researchers with patients’ experiences ^26^, and helps develops positive relationships between researchers and patients ^27^. Our findings indicate that preclinical spinal cord researchers and clinicians working in spinal cord injury appreciate the benefits of involving people affected by spinal cord injury can have for preclinical research. Barriers and facilitators identified by the participants may help inform the PPI activities of other preclinical research teams. While the perspectives of both groups were similar, their understanding of the role of PPI was based on their own professional backgrounds, which may lead to confusion or even conflict if these differences are not discussed. Training may help align diverse perspectives in advance of commencing PPI activities.

To the best of our knowledge, this is the first study to explore the perspectives of these two key stakeholder groups on PPI in preclinical research. A range of researchers and clinicians across career stages were included, so the findings capture a broad range of experiences. Limitations include that the majority of those interviewed were from the Republic of Ireland, so findings may not be transferrable to other jurisdictions, particularly those outside the USA, Canada, the UK, and Australia where PPI is less established in research. All participants were familiar with the concept of PPI, and several had prior experience, and while this was helpful in terms of their responses, this may not reflect the wider community of preclinical spinal cord researchers and clinicians. Further research would be needed to establish the broader perspectives of the spinal cord research community.

## Conclusions

Participants in this semi-structured interview study of preclinical spinal cord researchers and clinicians involved in spinal cord treatment recognised the value that PPI contributors can bring to preclinical research. They identified that PPI may have particular value in improving research relevance and motivating researchers who would otherwise not have contact with the people who may ultimately benefit from it. As key stakeholders in PPI in preclinical spinal cord research, they identified barriers and enablers that researchers should keep in mind, and highlighted the importance of building strong relationships with people affected by spinal cord injury for successful PPI. These insights can provide a basis for encouraging and facilitating best practices for PPI in preclinical spinal cord research and in preclinical research more broadly.

## Data Availability

All data produced in the present work are contained in the manuscript

## Notes

### Competing Interest Statement

The authors have declared no competing interest.

### Funding Statement

This study is conducted as part of a research collaboration funded by the Irish Rugby Football Union Charitable Trust (IRFU CT), Science Foundation Ireland Advanced Materials and BioEngineering Research Centre (SFI AMBER) and conducted by Tissue Engineering Research Group (TERG) at the Royal College of Surgeons in Ireland (RCSI) University of Medicine and Health Sciences. Padraig Carroll is also a recipient of a Clement Archer Postgraduate Scholarship from the RCSI School of Pharmacy and Biomolecular Sciences.

### Author Declarations

The Ethics Committee of the Royal College of Surgeons in Ireland, University of Medicine and Health Sciences gave ethical approval for this work

